# Sensitive detection and quantification of SARS-CoV-2 in saliva

**DOI:** 10.1101/2020.12.04.20241059

**Authors:** Mustafa Fatih Abasiyanik, Blake Flood, Jing Lin, Sefika Ozcan, Sherin J Rouhani, Athalia Pyzer, Jonathan Trujillo, Chaojie Zhen, Ping Wu, Stephen Jumic, Andrew Wang, Thomas F. Gajewski, Peng Wang, Madeline Hartley, Bekim Ameti, Rachael Niemiec, Marian Fernando, Bulent Aydogan, Cindy Bethel, Scott Matushek, Kathleen G. Beavis, Nishant Agrawal, Jeremy Segal, Savaş Tay, Evgeny Izumchenko

## Abstract

Saliva has significant advantages as a test medium for detection of SARS-CoV-2 infection in patients, such as ease of collection, minimal requirement of supplies and trained personnel, and safety. Comprehensive validation in a large cohort of prospectively collected specimens with unknown SARS-CoV-2 status should be performed to evaluate the potential and limitations of saliva-based testing. We developed a saliva-based testing pipeline for detection of SARS-CoV-2 nucleic acids using real-time reverse transcription PCR (RT-PCR) and droplet digital PCR (ddPCR) readouts, and measured samples from 137 outpatients tested at a curbside testing facility and 29 inpatients hospitalized for COVID-19. These measurements were compared to the nasal swab results for each patient performed by a certified microbiology laboratory. We found that our saliva testing positively detects 100% (RT-PCR) and 93.75% (ddPCR) of curbside patients that were identified as SARS-CoV-2 positive by the Emergency Use Authorization (EUA) certified nasal swab testing assay. Quantification of viral loads by ddPCR revealed an extremely wide range, with 1 million-fold difference between individual patients. Our results demonstrate for both community screening and hospital settings that saliva testing reliability is on par with that of the nasal swabs in detecting infected cases, and has potential for higher sensitivity when combined with ddPCR in detecting low-abundance viral loads that evade traditional testing methods.

## Introduction

Due to its efficiency to spread from people with mild or no symptoms, SARS-CoV-2 virus is highly contagious and has infected tens of millions of people around the world. Sensitive and specific detection of SARS-CoV-2 is crucial for containing its spread. Consistent with Centers for Disease Control (CDC) guidelines, most testing is currently performed by collecting upper respiratory specimens including nasopharyngeal (NP) and anterior nares nasal swabs (NS). Collection of upper respiratory swabs can cause complications in some patients^1-4^ and is associated with discomfort, which reduces compliance. Additionally, these swabs must be collected by a trained healthcare provider, which puts both the patient and healthcare worker at risk for infection. Requiring trained personnel also limits the ability to scale testing to higher volumes, especially in the regions hit hardest by the disease where healthcare staff are most burdened. Potential shortages of both swabs and personal protective equipment further make increasing the volume of swab tests difficult.

Several recent studies reported that SARS-CoV-2 is detectable in saliva, which provides a less invasive and more scalable specimen collection approach^5-7^. The ACE2 receptor that SARS-CoV-2 binds to and uses as main entry point into the cell is expressed in the oral mucosa, with particularly high expression on the tongue. Release of virus on aerosolized saliva droplets is an established mechanism for contact-free, long-distance viral transmission and makes detection of virus in the saliva particularly important^8^. Additionally, potential self-collection by the patient could help increase the scale of testing and allow new strategies like kit dissemination by mail. Another advantage of saliva is that it can provide a larger amount of test material than upper respiratory swabs, which may be the key to improving sensitivity.

To date, most of the studies attempting to detect SARS-CoV-2 in saliva have used limited case numbers, and often a direct comparison to gold-standard nasopharyngeal (NP) or nasal swab (NS) testing by a certified laboratory has been missing. While these shortcomings allowed a quick dissemination of results during a rapidly evolving pandemic, a comprehensive and methodical evaluation of saliva testing using a large sample cohort is required to fully assess its advantages and limitations. Recent studies evaluating suitability of saliva for COVID-19 testing revealed conflicting results. Initial proof-of-concept studies that collected saliva using a swab performed by a healthcare worker observed ∼ 85-90% sensitivity (concordance) when compared to NP swabs^9-11^. Self-collected saliva swabs demonstrated only 66% sensitivity in another study^9^. Studies that evaluated self-collection of saliva using the drool-technique (spitting into a sterile tube) revealed a range from 70 to 100% detection sensitivity compared to matched NP swabs^5-7,12,13^. One study that requested spitting into a tube found 77% sensitivity, while another study that required patients to keep spitting until a sterile cup was 1/3 full found 92% sensitivity. In short, although FDA has recently issued an Emergency Use Authorization (EUA) to a limited number of saliva detection methods, there is no consensus whether saliva can serve as a sensitive alternative test medium for the management of COVID-19 pandemic. As a result, almost all certified testing for the diagnostic detection of SARS-CoV-2 in patients is still performed by traditional nasal swab methods.

While the specimen source and collection approach are important aspects of clinical testing, so is the method of detection. Per CDC recommendations, most clinical laboratories currently detect the presence of virus in upper respiratory swab specimen by real-time reverse transcription polymerase chain reaction (RT-PCR). While this method has been rapidly implemented around the world as the gold standard for clinical diagnosis of SARS-CoV-2, detection and quantification of low amount of virus using RT-PCR is challenging. PCR is prone to inhibition when using complex samples, which can reduce assay efficiency and elicit false-negative results^14^. Most importantly, RT-PCR does not provide quantitative results without standard curves, which are not routinely performed by most diagnostic labs. When standard curve is performed, the differences in amplification efficiency between samples and standards can also introduce variation^15^. As viral load of SARS-CoV-2 in saliva and nasal swab samples tends to decrease over time^6,16,17^, a longitudinal quantitative assessment of changes in patient’s viral load could help clinicians make better treatment decisions, a task which is difficult with current RT-PCR methods.

Droplet digital PCR (ddPCR) is an ultra-sensitive method for direct nucleic acids quantification that uses water-oil emulsion droplet technology to fractionate a sample into ∼20,000 nanoliter-sized droplets, each containing zero or one target molecule. Counting the number of positive droplets results in absolute target quantification without the need for a standard^18^ or calibration curve^19^. Furthermore, by concentrating target molecules in smaller reactions, ddPCR can reduce the bias introduced by PCR inhibitors^20-22^ often found in clinical specimens^21,22^. While these properties make ddPCR a promising SARS-CoV-2 detection method for samples with low template abundance^20^, a comprehensive validation of this approach in a large cohort of prospectively collected specimens with unknown SARS-CoV-2 status has not yet been performed.

We therefore sought to evaluate ostensible improvements to current testing strategies. To this end, we have first developed, optimized and performed a thorough analytical validation of quantitative RT-PCR and ddPCR detection assays for SARS-CoV-2 in saliva using a range of positive and negative synthetic controls and clinically collected NP swabs with known SARS-CoV-2 status. We next used our RT-PCR and ddPCR assays to analyze NS and saliva specimens concurrently collected from 92 patients and 45 asymptomatic individuals at the curbside screening facility established at the University of Chicago Medical Center, and 29 additional inpatient cases with confirmed SARS-CoV-2 infection upon admission. An additional NS specimen was collected from each patient at the same time and analyzed in a CLIA-certified clinical microbiology laboratory (CML) using the FDA emergency use authorized (EUA) Cepheid Xpert Xpress SARS-CoV-2 test.

Our results demonstrate that saliva testing performed by either ddPCR or RT-PCR platforms shows over 90% sensitivity (concordance) for detecting positive cases identified by the diagnostic assay by CML. Particularly, quantification of saliva specimens by standard RT-PCR resulted in 100% agreement in identifying patients determined as SARS-CoV-2 positive by the certified CML nasal swab testing, showing excellent sensitivity. This result strongly supports the use of saliva testing as an alternative and practical method for detection of SARS-CoV-2 infection in patients. Furthermore, ddPCR saliva assay was able to detect additional positive cases that have been called as negative using the nasal-swab Cepheid Xpert Xpress test or by our own nasal-swab ddPCR testing. Quantification of curbside collected specimens revealed an extremely wide range of viral loads in patients, spanning a range of more than one million-fold. Furthermore, we observed overall higher viral loads in hospitalized inpatients’ saliva compared to the curbside collected samples. Collectively, our results provide a thorough evaluation of saliva as a clinical specimen for SARS-CoV-2 detection, supports saliva testing as a highly sensitive and feasible clinical diagnostic approach, and suggests potential for additional sensitivity when ddPCR readout is used.

## Results

### Validation of SARS-CoV-2 PCR assays with nucleic acid standards

We first optimized RT-PCR and ddPCR assays targeting nucleocapsid (N1) and nucleocapsid (N2) genes of SARS-CoV-2 using commercially available RNA standards. For both RT-PCR and ddPCR assays, we used one-step reverse transcription kits to simplify the PCR assays and to minimize errors. Our RT-PCR measurements showed excellent efficiency (>99.25 % for N1 and N2) and limit of detection (LOD; we were able to detect 1 copy of viral mRNA in a reaction). The RT-PCR cycle threshold (Ct) values corresponding to 1 copy number (CN) of N1 and N2 genes were calculated to be 38.2 and 38.6, respectively (**Supplementary Figure 1**). Similarly, the ddPCR assay targeting the N1 and N2 genes of SARS-CoV-2 showed excellent linearity and revealed an LOD of 0.06 and 0.21 CN/µL for N1 and N2, respectively (**Supplementary Figure 2**).

### Characterization of RT-PCR and digital-PCR sensitivity for detection of SARS-CoV-2

To characterize the sensitivity of both PCR assays in patient samples, nasal swab specimens collected from 35 COVID-19 negative individuals (as confirmed by the FDA EUA TaqPath COVID-19 kit and EUA Cepheid Xpert Xpress diagnostic systems, as well as research RT-PCR and ddPCR assays) were pooled together, mixed and aliquoted into individual vials. Nasal swab specimen collected from a clinically confirmed COVID-19 positive patient was then added to the pooled negative aliquots at different dilution ratios, ranging from 1/10^2^ to 1/10^7^ (**Supplemental Table 1**). Four identical dilution series were prepared in duplicates. One dilution set was tested with the Cepheid Xpert Xpress SARS-CoV-2 system. For the remaining three dilution sets, RNA was extracted using an automated KingFisher Flex instrument and analyzed using the FDA EUA Applied Biosystems TaqPath COVID-19 kit as well as RT-PCR and ddPCR assays optimized in our research laboratory settings. These assays have different positivity cut-offs. The diagnostic Cepheid Xpert Xpress system used by CML calls a NP or NS sample positive if either N2 or envelope (E) gene shows a Ct value lower than 45 ^23^. The TaqPath COVID-19 kit calls sample positive if any 2 of the 3 target genes [N gene (N1 and N2 regions), spike glycoprotein (S) gene and ORF1ab gene] show a Ct value lower than 37. The CDC guidelines however recommend calling an RT-PCR test positive when Ct values are lower than 40, and this threshold was used for our research laboratory RT-PCR assay. For ddPCR measurements in these diluted samples, a specimen was called positive when CN/µL for either N1 or N2 was higher than 0.1 and more than two positive droplets were detected in the reaction (as recommended by the manufacturer, Bio-Rad Inc.). While RT-PCR based tests (EUA Cepheid Xpert Xpress, EUA TaqPath and our own research laboratory RT-PCR assay) could reliably detect the presence of a virus in 1/100,000 diluted samples, ddPCR assay was able to detect N1 target gene at 1/1,000,000 dilution **(Supplementary Table 1)**. Although these results are suggestive of higher sensitivity of ddPCR platform, a direct comparison between assays using different RNA extraction methods, gene targets, cycling conditions, and cut-offs should be addressed with caution.

**Table 1:** Clinicopathological characteristics of the SARS-CoV-2 saliva testing participants.

| **Parameter – n (%)** | **Curbside (137)** | **Inpatients (29)** |
| --- | --- | --- |
| Age, median (IQR) - years | 45 (30,58) | 55 (41,65) |
| Male Sex - n (%) | 51 (37.2%) | 18 (62.1%) |
| **Race - n, (%)** |  |  |
| White | 56 (40.9%) | 8 (27.6%) |
| Black | 34 (24.8%) | 16 (55.2%) |
| Hispanic | 11 (8.0%) | 2 (6.9%) |
| Other | 6 (4.4%) | 1 (3.4%) |
| Unknown/declined | 30 (21.9%) | 2 (6.9%) |
| **Comorbidities per patient\* - n (%)** |  |  |
| None | 96 (70.1%) | 2 (6.9%) |
| 1 | 21 (15.3%) | 8 (27.6%) |
| 2 | 11 (8.0%) | 9 (31.0%) |
| 3 or greater | 9 (6.6%) | 10 (34.5%) |
| **Epidemiologic risk factors# - n (%)** |  |  |
| None | 44 (32.1%) | 1 (3.4%) |
| 1 | 43 (31.4%) | 3 (10.3%) |
| 2 | 31 (22.6%) | 9 (31.0%) |
| 3 or greater | 19 (13.9%) | 16 (55.2%) |
| **Medications at time of sampling - n (%)** |  |  |
| Hydroxychloroquine | 0 (0.0%) | 0 (0.0%) |
| Azithromycin | 0 (0.0%) | 10 (34.5%) |
| Lopinavir-Ritonavir | 0 (0.0%) | 0 (0.0%) |
| Systemic corticosteroid (within 24H of enrollment) | 0 (0.0%) | 1 (3.4%) |
| Remdesivir | 0 (0.0%) | 14 (48.3%) |
| **Supplemental O2 received - n (%)** |  |  |
| None | 0 (0.0%) | 8 (27.6%) |
| Low-Flow Oxygen | 0 (0.0%) | 13 (44.8%) |
| High-Flow-Oxygen | 0 (0.0%) | 5 (17.2%) |
| Invasive mechanical ventilation | 0 (0.0%) | 3 (10.3%) |
| **Inflammatory markers - at time of hospital admission, median (IQR)** |  |  |
| CRP - mg/L | NA | 67 (41-131) |
| D-dimer - pg/mL | NA | 1.26 (0.8-3.5) |
| Ferritin - pg/mL | NA | 744 (160-1792) |

\*Comorbidities were defined as heart failure, coronary artery disease, hyperlipidemia, history of cancer, systemic hypertension, pulmonary hypertension, home oxygen use, chronic lung disease, immunodeficiency, end stage renal disease, obesity, and diabetes.
#The risk factors were defined as being older than 60 years old, male, race (black, non-white Hispanic or Latino, American Indian, Alaskan native), presence of underlying comorbidity, and being immunocompromised.

We next assessed the sensitivity of the RT-PCR and ddPCR assays (both use the same primers and probes combination) in a cohort of 8 positive and 18 negative NP samples (based on CML results) from patients undergoing clinical testing for suspected COVID-19. For the positive samples we saw 100% concordance of RT-PCR and ddPCR results with CML data (i.e. all positive samples by CML were also positives in our measurements, **Supplementary Figure 4**), but out of the 18 samples called negative by the CML, 11 samples were identified as positive by RT-PCR and 13 by ddPCR. These additional positive cases were mostly samples with low viral load, suggesting a higher detection sensitivity of our optimized assays. Both RT-PCR and ddPCR assays have also demonstrated a high agreement in detection of an internal control RnaseP gene (**Supplementary Figure 4**), indicating that detection of additional positive cases by ddPCR is not biased by the input material. These assays were then used for further measurements on additional patient samples (N=137) and compared to NS testing, as described below.

### Curbside sample collection pipeline at The University of Chicago Medical Center

We established a sample collection pipeline from the clinical curbside testing site at the University of Chicago Medical Center. The pipeline was divided into 3 stages: samples collection at the curbside testing clinic, RNA extraction at a nearby BSL3 facility, and analysis with RT-PCR and ddPCR in a BSL1 research laboratory (**Figure 1**). We simultaneously collected three samples (2 NS and 1 saliva sample) from 137 volunteers (**Table 1**) who were scheduled at the curbside testing site to receive a standard-of-care clinical COVID test. 92 of these patients were being tested for COVID-19 related symptoms, and 45 asymptomatic subjects were being tested as screening prior to a planned medical procedure. Upon collection, one NS sample was sent to CML for clinical diagnostic analysis. The other NS and saliva samples were sent to a BSL3 facility for viral RNA extraction (see Methods). The extracted viral RNA samples were then transported to a BSL1 lab for concurrent RT-PCR and ddPCR analysis (**Figure 1**).

**Figure 1:**
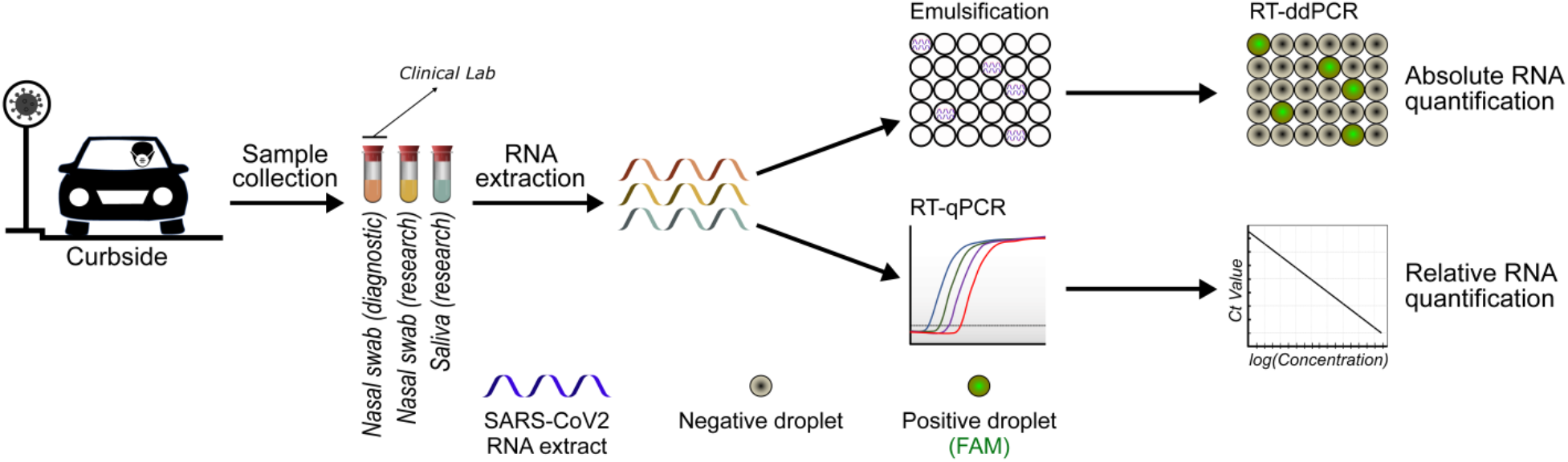
COVID-19 curbside patient sample collection and testing pipeline. Saliva and matched nasal swab (NS) specimens were simultaneously collected from 137 outpatients scheduled for SARS-CoV-2 testing at the University of Chicago curbside facility. This cohort included 92 individuals that were tested due experiencing symptoms of influenza-like illness, and 45 asymptomatic patients that were tested as part of routine pre-operative screening. Samples were also collected from 29 inpatients hospitalized for COVID-19. Upon sample collection, one NS sample was tested in the clinical microbiology laboratory (CML) using an FDA-EUA Cepheid Xpert Xpress kit by RT-PCR, which targets N1 and E genes of SARS-CoV-2. For the research samples, RNA was extracted in a BSL3 facility, and the eluates were analyzed with RT-PCR and droplet digital PCR (ddPCR) in the research laboratory. N1, N2 and RnaseP genes of SARS-CoV-2 were targeted by specific TaqMan probes with both assays. The concentrations of targets were calculated by using positive standards in each RT-PCR run.

### TRIzol-based RNA extraction for SARS-CoV-2 testing

During the height of the COVID-19 pandemic, many laboratory supplies such as RNA extraction kits were either not available or backordered. We therefore tested if a TRIzol-based RNA extraction (TRE) method, a widely used and simple approach for RNA extraction in research laboratories, would be feasible for COVID-19 screening tests. RNA extracted from the same saliva sample using either TRE or two CDC recommended commercial viral extraction kits ^24^ (QIAmp MinElute Virus DNA/RNA Spin Kit [QVDRK] and QIAmp Viral RNA [QVK] Mini Kit) was analyzed with RT-PCR and ddPCR to quantify the RnaseP gene, an internal control for extraction procedure. The relative efficiency of TRE method compared to QVK was found to be 17% (**Supplementary Figure 3**). Although this value is relatively low, the high sensitivity of our PCR assays compensates for the reduced extraction efficiency, and the availability of the reagents was considered a major advantage. We therefore proceeded to use this simple and available RNA extraction method for our studies.

### High concordance of saliva testing with clinical nasal swab assay

We measured the curbside collected patient specimens using our RT-PCR and ddPCR assays. As an assay control, a serial dilution of SARS-CoV-2 RNA standards was always analyzed along each RT-PCR or ddPCR plate (**Supplementary Figure 6 and 7**). The distributions of both RT-PCR and ddPCR results demonstrated an excellent concordance between N1 and N2 detection (**Figure 2a, b**). All 16 patient samples detected as SARS-CoV-2 positive by the CML were also called positive by our saliva RT-PCR test (**Figure 2c, d**), resulting in 100% concordance, and demonstrating the applicability of using saliva for sensitive SARS-CoV-2 diagnostic testing. Furthermore, our saliva testing detected an additional 7 positive cases that were called SARS-CoV-2 negative by the CML. Eighteen paired NS specimens collected from the 23 patients with positive saliva samples were also tested positive using the same research RT-PCR assay.

**Figure 2:**
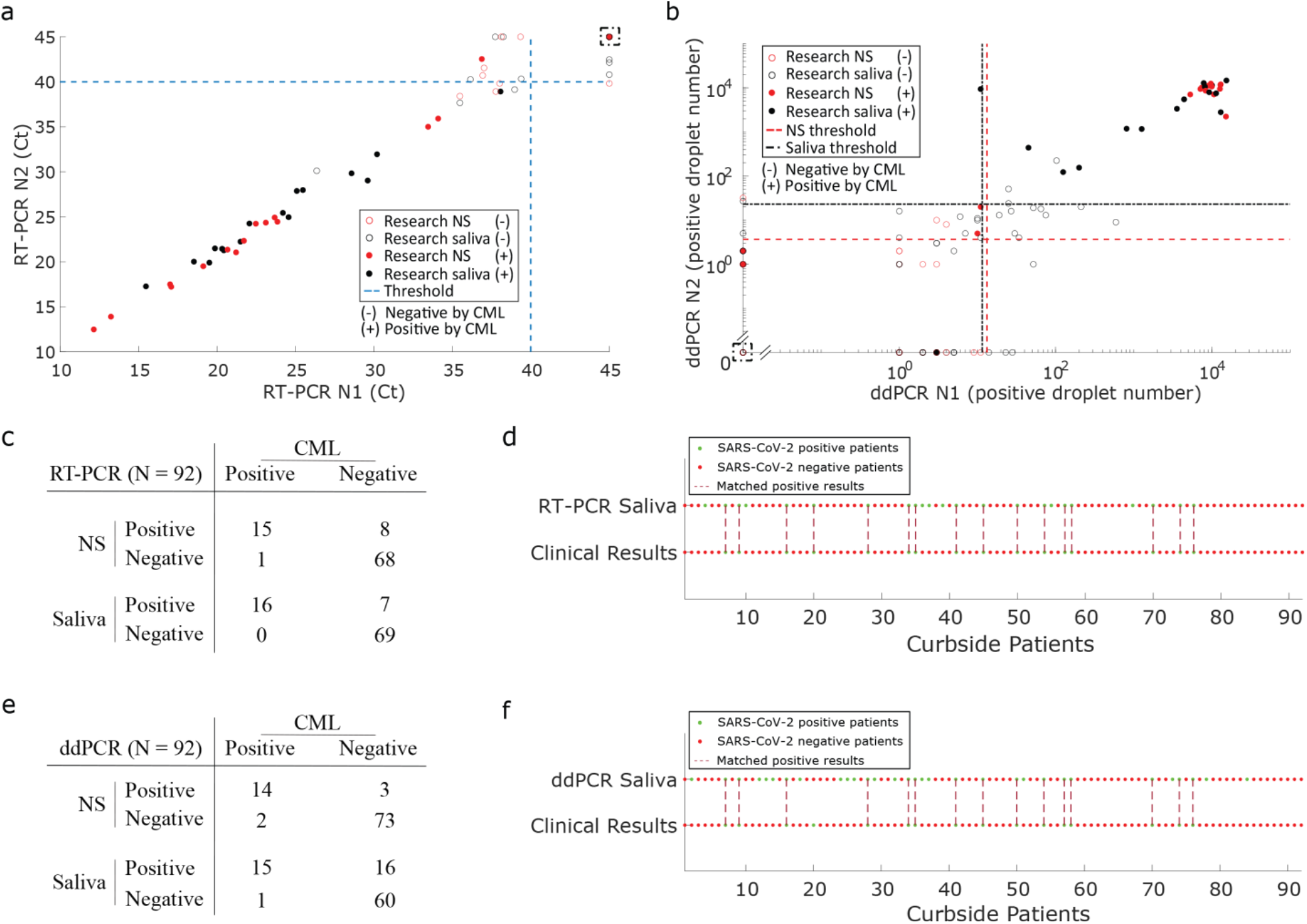
Saliva-based SARS-CoV-2 testing shows excellent concordance and sensitivity with nasal swab assay across curbside collected patient specimens. Curbside collected NS samples were analyzed at the certified microbiology laboratory (CML) using a RT-PCR based EUA approved assay, while matched saliva samples and additional NS specimen collected from each individual were analyzed by our research laboratory using optimized RT-PCR and ddPCR assays. (**a**) N1 vs. N2 gene concordance for NS and saliva samples measured by the RT-PCR assay at our research laboratory. Filled dots represent cases tested positive by the EUA approved diagnostic assay. Dashed blue lines at Ct value of 40 for N1 and N2 indicate CDC-recommended thresholds for RT-PCR assays using these primers/probes. Dashed box contains data from multiple samples with Ct = 45 for N1 and N2 (a total of 67 NS and 66 saliva samples). (**b**) N1 vs. N2 gene concordance for NS and saliva samples measured by the ddPCR assay at our research laboratory. The filled dots represent cases tested positive by the EUA approved diagnostic assay. The dashed lines indicate ddPCR thresholds determined by analyzing a cohort of 45 asymptomatic individuals who were SARS-CoV-2 negative by the CML test (**Supplementary Table 2**). Dashed box includes samples with 0 positive ddPCR droplets for N1 and N2 (a total of 38 NS and 17 saliva samples). (**c**) Comparison of cases detected by our NS and saliva RT-PCR assays to the CML NS results. Each number in the table represents cases detected as positive or negative by both our RT-PCR assay and by the CML. For example, in NS samples our RT-PCR assay detected 15 positive patients that are also detected as positive by the CML. On the other hand, our NS RT-PCR assay detected additional 8 positive patients that are detected as negative by the CML. Our saliva RT-PCR assay detected all (16 out of 16) positive cases that were detected as positives by the CML NS test. (**d**) Comparison between matched research RT-PCR saliva results and diagnostic facility NS results. The green and red dots indicate positive and negative cases respectively. The maroon dashed lines connecting the dots indicate patients tested positive by both assays. (**e**) Summary of the positive cases detected in NS and saliva samples by ddPCR assay (as in **c**). (**f**) Comparison between matched ddPCR saliva results and diagnostic facility NS results. The green and red dots indicate positive and negative cases respectively. The maroon vertical dashed lines connecting the dots indicate patients tested positive by both assays.

To evaluate the concordance of our ddPCR saliva assays with NS results from a clinical laboratory, we first sought to determine the baseline levels of SARS-CoV-2 nucleic acid signals in COVID-19 negative patients. Since no FDA recommended guidelines are available for ddPCR positive calling criteria, and because manufacturers recommendations (BioRad) were provided for nasal swab testing only, we have developed the thresholds for both N1 and N2 genes *de novo* using specimens collected from healthy individuals in our pre-operation screening cohort (pre-op). This cohort contained 45 asymptomatic individuals who participated in the curbside testing as part of their pre-operative screening process. All of these patients were SARS-CoV-2 negative by CML nasal swab test. Using a standard formula for threshold detection (mean+2*SD, SD: standard deviation), the new ddPCR saliva thresholds were calculated and listed in **Supplementary Table 2**. Data distribution of the healthy cohort is shown in **Supplementary Figure 5**.

We have next applied these empirical ddPCR thresholds to determine the COVID-19 status of the remaining 92 subjects collected at the curbside (**Figure 3**). A patient’s sample was called positive when either N1 or N2 target genes showed both CN/µL (concentration) and positive droplet number above the respective thresholds (see Methods). We found that ddPCR results generated from saliva specimens in our research lab were highly concordant with the diagnostic data obtained from the nasal swab (NS) test performed at the certified CML. Our saliva ddPCR testing identified 15 out of 16 individuals determined as positive by the NS-based CML assay (**Figure 2e, f**), achieving a concordance of 93.75% in detecting positive cases. In matched NS specimens, ddPCR assay has identified 17 positive cases, including 14 cases called positive by the CML (**Figure 2e**).

**Figure 3:**
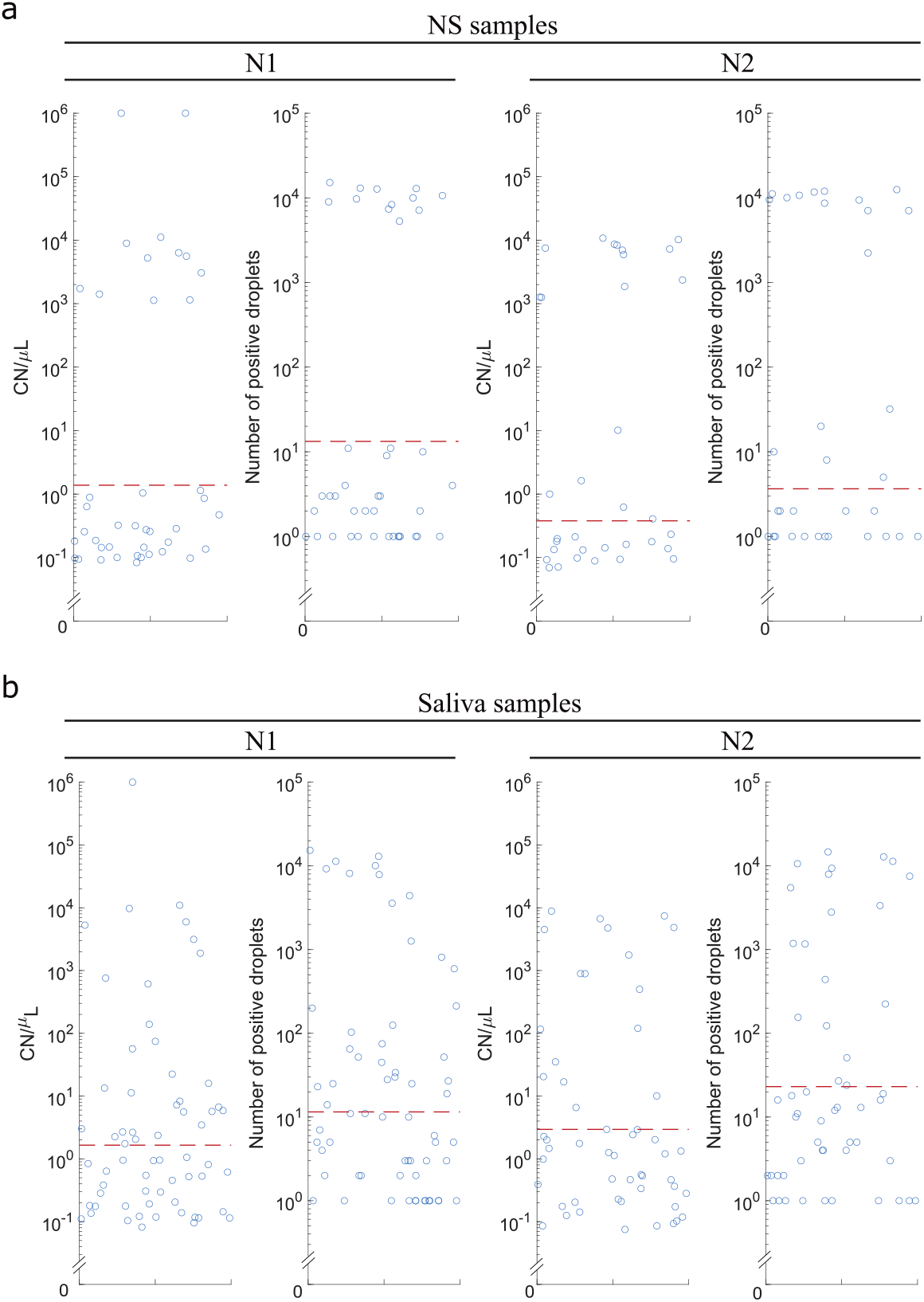
Quantification of SARS-CoV-2 load in NS and saliva samples by ddPCR. Each dot represents ddPCR results from distinct nasal swab (**a**) or saliva specimen (**b**) collected from 92 patients with reported symptoms at the curbside testing facility. Red dashed lines indicate ddPCR thresholds calculated using measurements from 45 SARS-CoV-2 negative and asymptomatic individuals (pre-operative cohort). Droplets with concentration/counts above the threshold were determined as SARS-CoV-2 positive.

We next analyzed an independent specimen cohort collected from 29 inpatients admitted to the hospital with COVID-19. For these subjects, NS and saliva were collected simultaneously. NS samples were tested by the CML, whereas saliva specimens were tested by our ddPCR and RT-PCR assays in the research laboratory. Important to note, while all inpatients were positive by CML upon diagnosis, NS and saliva samples for research purposes were collected several days after patients have been already admitted to the hospital. Of the 29 NS samples, 14 were called SARS-CoV-2 positive by the CML. ddPCR analysis of the matched saliva specimens detected 17 positive cases, including 11 subjects identified by the CML. Saliva analysis by RT-PCR resulted in 23 positive cases, including 14 subjects that were positive by CML, indicating higher detection sensitivities with ddPCR and RT-PCR saliva assays (**Figure 4**).

**Figure 4:**
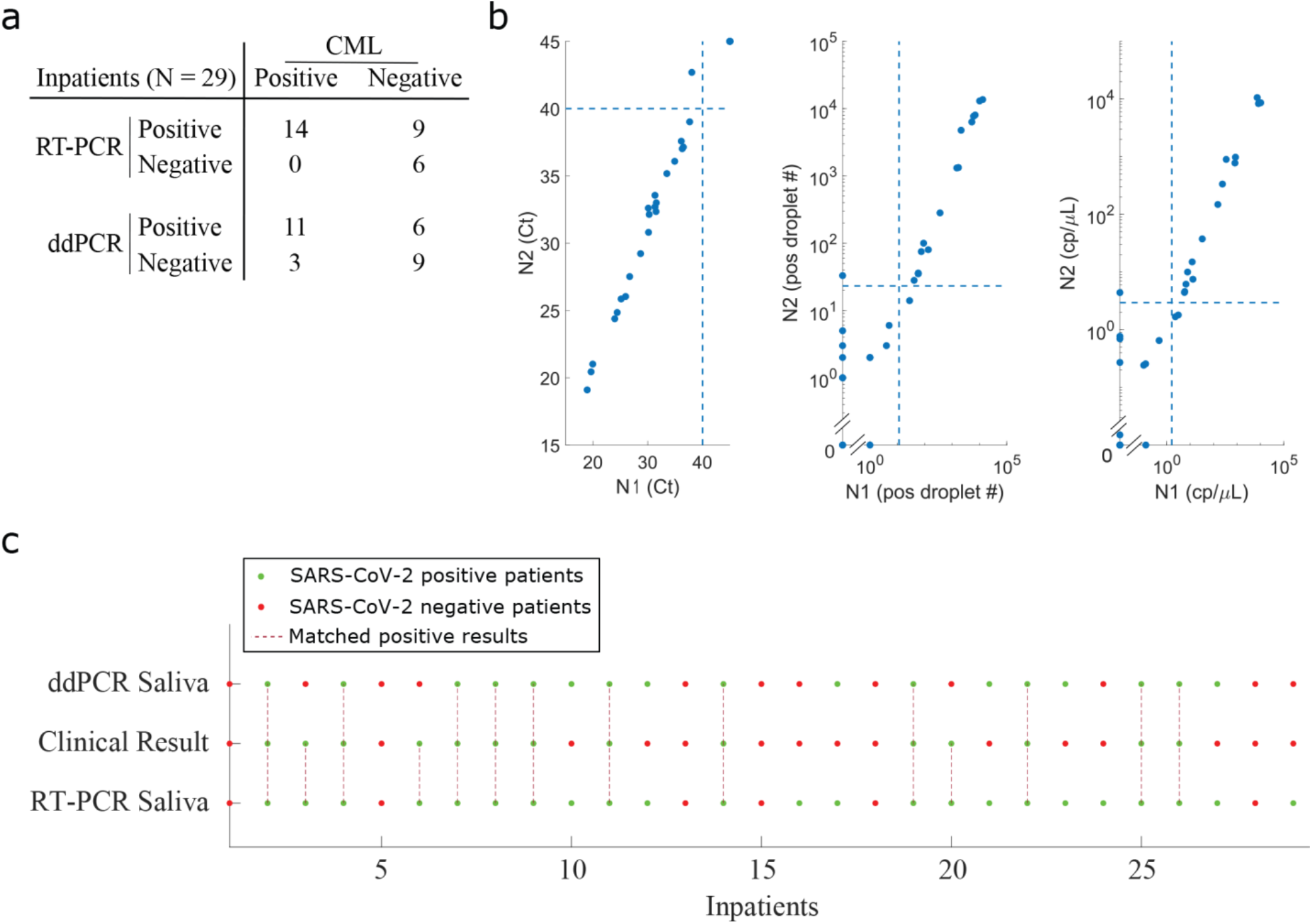
Analysis of nasal swab and saliva samples collected from 29 inpatients hospitalized for COVID-19. (**a**) Table summarizes results of our research-based RT-PCR and ddPCR assays compared to CML nasal swab testing. Each number in the table represents cases detected as positive or negative by our assays and by the CML. For example, our saliva RT-PCR assay detected all (14 out of 14) positive patients that are detected as positive by the CML. On the other hand, our RT-PCR saliva assay detected an additional 9 positive patients that are deemed as negative by the CML. (**b**) The figure demonstrates N1 vs N2 target gene concordance assessed by RT-PCR Ct values (left), ddPCR positive droplet number (middle), and ddPCR copy number per µL (right). The dashed lines in each subplot indicate empirical thresholds for positive readout calling. (**c**) The comparison of matched saliva-based RT-PCR and ddPCR results with NS-based results generated by the diagnostic facility. For saliva ddPCR assay, a sample is called positive if either N1 or N2 are above the empirical thresholds. For saliva-based RT-PCR assay, a sample is called positive if either N1 or N2 show Ct values < 40. The green dots indicate positive sample status, while red dots indicate samples tested negative in respective settings. The maroon dashed lines indicate samples tested positive by the respective assays.

### Potential for increased sensitivity in detecting SARS-CoV-2 with saliva digital-PCR readout

Overall, by analyzing saliva samples we were able to identify most curbside subjects that were determined as positives by the CML (100% for RT-PCR, 93.75% for ddPCR assays), (**Figure 2c-f**), demonstrating the applicability of using saliva for sensitive SARS-CoV-2 diagnostic testing. While NS-based CML assay has detected 16 of the 92 cases as positives for SARS-CoV-2, ddPCR assay identified 17 and 31 positives from NS and saliva samples respectively (**Figure 5**). Due to a higher sensitivity and resolution provided by the ddPCR assay, we were able to catch more positive cases than RT-PCR (8 more) and CML (15 more), especially in cases where the viral copy number is very low (**Figure 2a, b, Figure 3 and Figure 5**). This is especially noticeable for saliva samples assessed with ddPCR (black circles in **Figure 2b**). The ability to detect low viral load in patients may be useful for identifying potential viral carriers with no or mild symptoms.

**Figure 5:**
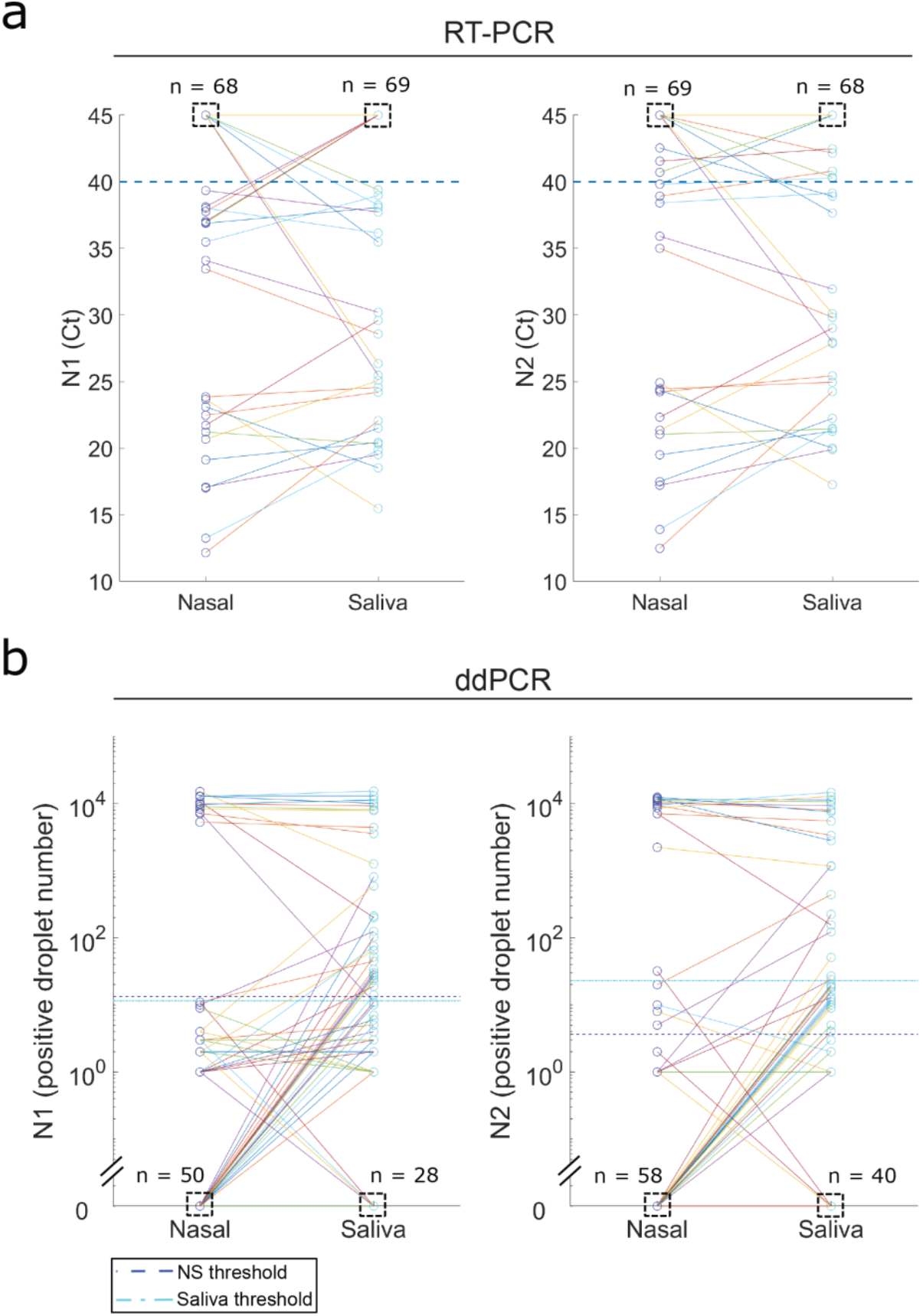
Comparison of research RT-PCR (a) and ddPCR (b) results between nasal swab and saliva. (**a**) Dashed boxes indicate number of negative samples with Ct value of 45. The number of nasal swab specimens with Ct value of 45 for N1 and N2 was 68 and 69 respectively, whereas the number of saliva samples with Ct value of 45 for N1 and N2 was 69 and 68 respectively. There were 9 and 7 nasal swab samples that were negative for N1 and N2 respectively, but these patients were detected as positive for these targets in matched saliva samples. There were also 3 and 3 nasal swab samples that were positive for N1 and N2 respectively, but called negative for these targets in matched saliva specimens. Per CDC recommendations, a sample is called positive if either N1 or N2 are below the RT-PCR thresholds. (**b**) Dashed boxes indicate number of samples with readout of 0. The total number of negative samples with concentration readout of 0 for N1 in nasal swab and saliva specimens was 50 and 58 respectively, while the number of negative samples with concentration readout of 0 for N2 in nasal swab and saliva specimens was 28 and 40 respectively. The number of nasal swab negative but saliva positive results are 19 (N1) and 5 (N2) and the number of nasal swab positive but saliva negative results are 1 (N1) and 3 (N2).

To confirm that additional samples detected by ddPCR are true positives and verify whether the readings remain consistent and reproducible over time, we independently re-analyzed remnant RNA specimens extracted from 46 saliva samples (19 positive and 27 negative, **Supplementary Table 4**) with ddPCR. The samples selected prioritized individuals that were caught positive only by ddPCR but missed by CML or our RT-PCR tests. The results of two independent analyses showed excellent consistency (**Supplementary Figure 8**). Out of the 46 re-tested samples, 19 were indicated as positives during the first run, of which 15 were confirmed as positives in the second run. The 4 samples with disagreement between the two runs had PCR readings close to our empirical thresholds. While some specimens showed lower readings upon re-analysis compared to first ddPCR run, this may be attributed to the RNA degradation after storage for nearly 5 months. All 27 samples that were negative during the first run were confirmed to be negative upon re-analysis.

## Discussion

An important step in containing the spread of COVID-19 is early detection of viral carriers in public and monitoring viral load changes in patients. Conventional sample collection methods such as nasal or nasopharyngeal swabs are invasive, time consuming, labor intensive, and rely on availability of nasal swabs which can be in short supply. In addition, these methods increase the infection risk to health care workers by provoking patients to sneeze and cough^25^. Nasal swab (NS) testing is uncomfortable for patients^26^, possibly leading to low adherence. Saliva testing offers a non-invasive, easy to collect alternative method that could reach a wider range of the population with reduced cost and risk to healthcare workers. Despite the potential of saliva testing, however, previous studies revealed conflicting results on its sensitivity for detection of SARS-CoV-2 in patient specimens and for its concordance with, FDA EUA approved nasal swab testing by clinical laboratories.

To this end, we have compared two different sampling approaches (saliva and NS) by using readouts from RT-PCR and ddPCR methods as well as an EUA approved diagnostic platform, demonstrating the benefits of saliva as a test medium for detection of SARS-CoV-2 infection in patients. We first demonstrate that our optimized ddPCR and RT-PCR assays can respectively detect virus in a positive NP sample diluted at 100,000 and 10,000 times in a pool of negative specimens, respectively (**Supplementary Table 1**). The high sensitivity of detection provides support for a suitability of the ddPCR platform for future pooled COVID-19 screening approaches. Analysis of matched saliva and NS specimens demonstrate a very high concordance between saliva-based assays and clinical nasal swab testing in determining positive cases. The clinical microbiology laboratory detected 16 positive patients out of 92 samples collected at the curbside facility. Out of 16 clinically positive patients, 16 (100%) and 15 (93.75%) were detected as positive by our saliva-based RT-PCR and ddPCR readout, respectively. This high concordance in detecting the positive patients with clinical diagnostic testing further supports the suitability of saliva testing as a noninvasive alternative method for the detection of SARS-CoV-2 in patients.

ddPCR measurements on saliva specimens revealed a higher number of positive patients compared to nasal swabs performed by the approved diagnostic laboratory. Out of 92 curbside collected patient specimens, saliva testing detected 31 positive samples with ddPCR readouts, a substantially higher number than that detected by the clinical diagnostic platform using NS specimens or our own NS ddPCR measurements. These additional positive calls were close to but above the threshold of our assay and were seen in patients with symptoms. Notably, independent re-testing of 46 saliva specimens with ddPCR confirmed presence of SARS-CoV-2 with 91.3% concordance, supporting a high reproducibility of this screening approach. In line with these observations, a comparable higher sensitivity was also obtained by Whyllie et al, using a RT-PCR saliva based readout^7^. Similarly, Suo et al. reported that 26 patients deemed negative by RT-PCR (out of 77 patients) were detected as positive by ddPCR^27^. In another study, multiplex ddPCR was used to test saliva and NP samples from 130 individuals with COVID-19 symptoms, and ddPCR was found to be superior to RT-PCR in determining positive cases^28^. Results published by several recent studies also support the use of ddPCR as a sensitive method for SARS-CoV-2 detection and quantification^29-31^.

Our analysis also revealed a very wide range in viral load measurements detected among COVID-19 positive patients, with up to 10^6^ fold range in viral load for both curbside patients and inpatients (**Supplementary Table 3**). Interestingly, some patients showed extremely high viral nucleic acid concentrations, over 10^6^ CN/ml. Quantification of viral load enabled us to classify these patients as supercarriers (viral loads>10,000 CN/ml sample), high viral level carriers (between 1,000 and 10,000 CN/ml sample), moderate viral level carriers (between 100 and 1,000 CN/ml sample), and low viral level carriers (<100 CN/ml sample). We found that saliva measurements show a more uniform distribution of viral loads among patients compared to nasal swabs. While most of the nasal swab samples showed either very high or very low viral loads, saliva testing revealed a more gradual distribution of viral loads in the same patients, indicating that saliva may be a more representative test medium for quantification of the overall viral load in a patient. The viral load detected by NS may also be influenced by how aggressively the swab is done, whereas there may be less operator-variability in collecting an equivalent volume of saliva. The precise quantification of viral load in saliva is crucial for identifying patients with high potential for transmission, as saliva is a main source of larger droplets and aerosols that are believed to spread the virus^32^. Recent evidence suggests a close relationship between patients with high viral load (supercarriers) and superspreaders^33^, which makes the saliva among the most relevant human specimens to determine superspreaders^29-32^

In summary, our work demonstrates the utility and sensitivity of saliva-based testing for detection of SARS-CoV-2 in a large group of patients from different settings (healthy, curbside, inpatients), and provides a comprehensive characterization of this method by comparing it to nasal swab testing performed by a clinical microbiology laboratory. We showed that measurement of saliva specimens provides high concordance with nasal swab testing, with possibility to provide additional sensitivity in detecting patients with low viral load. The inherent advantages, sensitivity, scalability and safety of saliva testing, makes it an important alternative to nasal or nasopharyngeal swab-based assays. While we believe that saliva testing is already a feasible alternative to traditional methods and should be immediately utilized, we also believe that improvements in pooled testing methods, automation, and parallelization will enable a much wider, rapid and cost-effective application of this important testing method in the management of the COVID-19 pandemic.

## Data Availability

Data is available upon request from the corresponding authors.

## Acknowledgements

We acknowledge discussions with BioRad, Inc. for guidance in ddPCR threshold calculations. We thank Sarah Kundrat and Melissa Gray for their role in patient recruitment and sample collection.

## Funding

This study was supported in part by U.S. National Institutes of Health (NIH) grants R01GM127527 to S.T., R01DE027809 to E. I., and R01DE028674 to N. A., and Paul. G. Allen Distinguished Investigator Award to S.T. We thank The University of Chicago Pritzker School of Molecular Engineering, and The University of Chicago Biological Sciences Division, for supporting this study.

## Conflicts of Interest

The authors declare no conflicting interests.

## Authors’ Contributions

E.I., S.T., J. S., N.A., M.F.A. conceived the project; S.O., P.W., R.N., M.F.A., J.L., M.F., C.B., M.S., A.W., R.N., B.F., S.J.R., A.P., J.T., B.A., C.Z., M.H., B.A. performed experiments and analyzed the data; E.I., N.A., S.T., J.S., T.F.G. acquired project funding; S.T., N.A., E.I., J. S., M.F.A. developed methods; S.T., J. S., N.A., E.I., T.F.G., K.G.B. provided experimental resources; S.J., B.F., C.Z., A.W., P.W., M.F.A., J.L., M.F., J.S., E.I. curated the data; J.L., M.F.A., E.I., S.T. created figures and tables; E.I., S.T., N.A., J. S., T.F.G., K.G.B. supervised the project; M.F.A., J.L., E.I., S.T., B.F., J.S. wrote the original draft; all authors reviewed and edited the final manuscript. E.I., S.T. J.S. M.F.A. verified the underlying data.

## Ethics Committee Approval

This study was approved by the Institutional Review Board (IRB) of the University of Chicago (IRB 20-0520), and informed written consent was obtained from all patients before sample collection.

## Methods

### Sample collection

This study was approved by the Institutional Review Board (IRB) of the University of Chicago (IRB 20-0520), and we obtained informed written consent from all patients prior to sample collection. 137 outpatients who were scheduled for clinical curbside testing either due to the presence of influenza-like symptoms (n = 92) or as part of routine pre-operative or pre-procedural screening (n = 45) agreed to participate. Enrolled patients received a clinical COVID-19 test administered by the clinical team, followed by an additional nasal swab and saliva specimen collection administered by the research team. Paired NS and saliva samples were also collected from 29 inpatients admitted with COVID-19. The saliva sample was collected by instructing patients to spit until they had filled a sterile tube to the 2 mL line and then 2 mL of Zymo DNA/RNA shield solution (ZymoR1100-50) was immediately added. NS samples were collected into a tube containing 1mL of Amies transport solution (Thermo Scientific).

### RNA extraction

For QIAmp MinElute Virus DNA/RNA Spin Kit (QVDRK) (Qiagen) and QIAmp Viral RNA Mini Kit (QVK)) (Qiagen) RNA extraction was performed following the manufacturer’s instructions. For Trizol™ LS Reagent (Thermo Fisher Scientific, #10296010) the manufacturer’s protocol was modified as follows: 1 ml of sample (i.e., saliva or nasal swab), 3 ml of Trizol LS Reagent and 0.6 ml of chloroform were mixed and centrifuged for 15 min at 12,000 x g at 4°C. 0.8 ml of the colorless upper aqueous phase was carefully transferred to a new tube containing 1 ml isopropanol and 1 µl glycogen. The mixture was kept at -20 °C overnight or was kept at room temperature for 10 min. After that, the mixture was span for 10 min at 12,000 x g, 4°C, and the supernatant was discarded. The pellet was resuspended in 2 ml 75% ethanol, vortexed mixing and centrifuged for 5 min at 7500g, 4°C. The supernatant was discarded, and the pellet was incubated at room temperature for 5-10 minutes. After ethanol is evaporated, the pellet was resuspended in 50 µl RNase-free water, the quantity and quality of extraction was assessed using Qubit RNA HS kit, and stored at -80 °C.

### ddPCR assay

The 20 μl ddPCR reaction containing 5 µl One-step ddPCR Supermix, 2 µl reverse transcriptase, 1 µl 300 mM DTT (Bio-Rad), 2 µL N1/N2/RnaseP probe/primers sets (IDT, #10006770), and 6.4 µl of sample were loaded into either manual or automated droplet generator (Bio-Rad). The emulsion was transferred into a 96 well plate, sealed, and cycled using a C-1000 thermal cycler (Bio-Rad) under the following conditions: 25°C for 2 min, 50°C for 60 min, 95°C for 10 min, 45 cycles of 95°C for 30 seconds, 55°C for 1 min, then 98°C for 10 min, and 4°C for infinite hold (ramping speed was 2.5°C/s). After amplification, the plate was transferred to a QX200 droplet reader (Bio-Rad) from which raw fluorescence amplitude data is extracted to the Quantasoft software for downstream analysis. NS samples were called positive when CN/µL for either N1 or N2 was higher than 0.1 and more than two positive droplets were detected in the reaction, according to the manufacturer recommendation. As no Bio-Rad recommended threshold was available for saliva specimens, an empirical threshold was set by collecting and analyzing samples from 45 asymptomatic participants. All these individuals were COVID-19 negative by CML. A standard formula: mean+2*SD (standard deviation), was utilized for the determination of a threshold. The thresholds ae reported in **Supplementary Table 2**.

### RT-PCR assay

The 20 µl RT-PCR reaction mixture contained 5 µl TaqPath™ One-Step RT-PCR Master Mix (Thermo Fisher Scientific), 2 µL of N1, N2 or RnaseP probe/primers sets (IDT 2019-nCov CDC EUA, IDT, #10006770), 0.1 µl Precision Blue Real-Time PCR Dye (Bio-Rad,), 6.4 µL of sample and 6.5 µL of PCR grade water. 2019-nCoV_N_Positive Control (IDT, #10006625) was used as DNA-based positive control for the DNA amplification part of the reaction. SARS-CoV-2 RNA Standard (Exact Diagnostics, #COV019) was used as a RNA-based positive control. Millipore nuclease free water processed with RNA extraction methods was used as negative control. IDT nuclease free water was used as a non-template control. CFX384 Touch Real-time PCR detection system (Bio-Rad,) was programmed as follows based on CDC’s recommendations^24^: 25°C for 2 min, 50°C for 15 min, 95°C for 2 min, 45 cycles of 95°C for 3 seconds, 55°C for 30 s. The results were evaluated by CFX Maestro™ Software (Bio-Rad). NS and NP samples were called positive when Ct values for either N1 or N2 targets were lower than 40, as recommended by the CDC. The same criteria were used for analysis of saliva specimens.

### TaqPath COVID-19 assay

RNA was extracted with MagMAX viral/pathogen (ThermoFisher) nucleic acid isolation kit using KingFisher Flex instrument for automated sample purification. Extracted RNA was reverse transcribed to cDNA and amplified/detected using the EUA TaqPath COVID-19 Combo Kit (ThermoFisher) on an Applied Biosystems 7500 Fast RT-PCR instrument. The assay contains probes to detect and amplify three SARS-CoV-2 specific target genes: ORF1ab, N gene, and S gene. If three targets are detected or if any 2 of the 3 target genes are detected, the test reports a positive result.

### Cepheid Xpert Xpress SARS-CoV-2 test

The GeneXpert Dx system (Cepheid) is an integrated diagnostic device that performs automated specimen processing and real-time RT-PCR analysis. The Xpert Xpress test consists of two main components: (i) the plastic cartridge, which contains liquid sample-processing and PCR buffers and lyophilized real-time RT-PCR reagents, and (ii) the GeneXpert Dx system, which controls intracartridge fluidics and performs real-time RT-PCR analysis. The EUA approved Xpert Xpress test was designed to amplify sequences of the envelope (E), nucleocapsid (N2), and RNA-dependent RNA polymerase genes. Only results from the E and N2 targets are used to generate test results. If both targets are detected, or if only N2 is detected, the test reports a positive result. If only the E target is detected the test reports a presumptive positive result because the target is shared among some members of the Sarbecovirus subgenus of coronaviruses. The EUA approved Xpert test allows the user to see amplification curves and Ct values for the N2 and E targets only.

### Statistical analysis

All figures were plotted with MATLAB or R software. Relative concentrations for N1, N2, and RnaseP targets assessed by the RT-PCR assay were estimated using linear regression analysis of dilution series of the RNA standard (SARS-CoV-2 RNA Standard, Exact Diagnostics).

## Supplementary materials

**Supplementary Figure 1:**
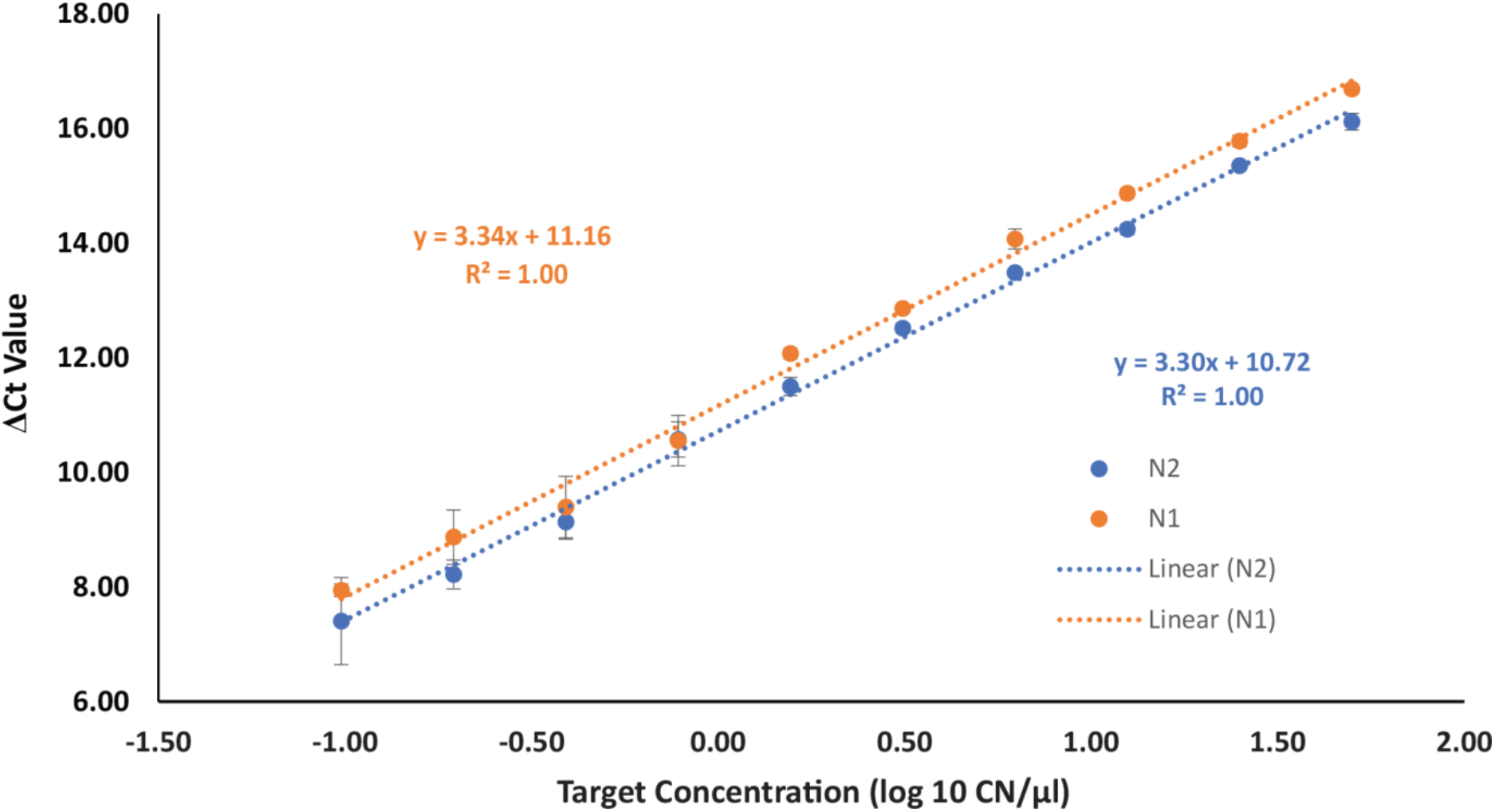
RT-PCR calibration curves for N1 and N2 targets. Serial dilution of N1 and N2 standards showed calibration curves with high concordance and linearity. The RT-PCR efficiency for each target are greater than 99.25 %. The lowest standard concentration used (i.e., 0.1CN/µl, CN: copy number) can be easily discriminated from negative template control (whose Ct value was 45). The technical limit of the assay is 1 CN of target in a reaction mixture of 20 µL, with corresponding Ct cycles (according to our calculated slope and intercept) are 38.2 and 38.6 cycles for N1 and N2 respectfully. We used the CDC guided Ct threshold (i.e., <40) to call a sample positive (n=3, the error bars: Standard Error).

**Supplementary Figure 2:**
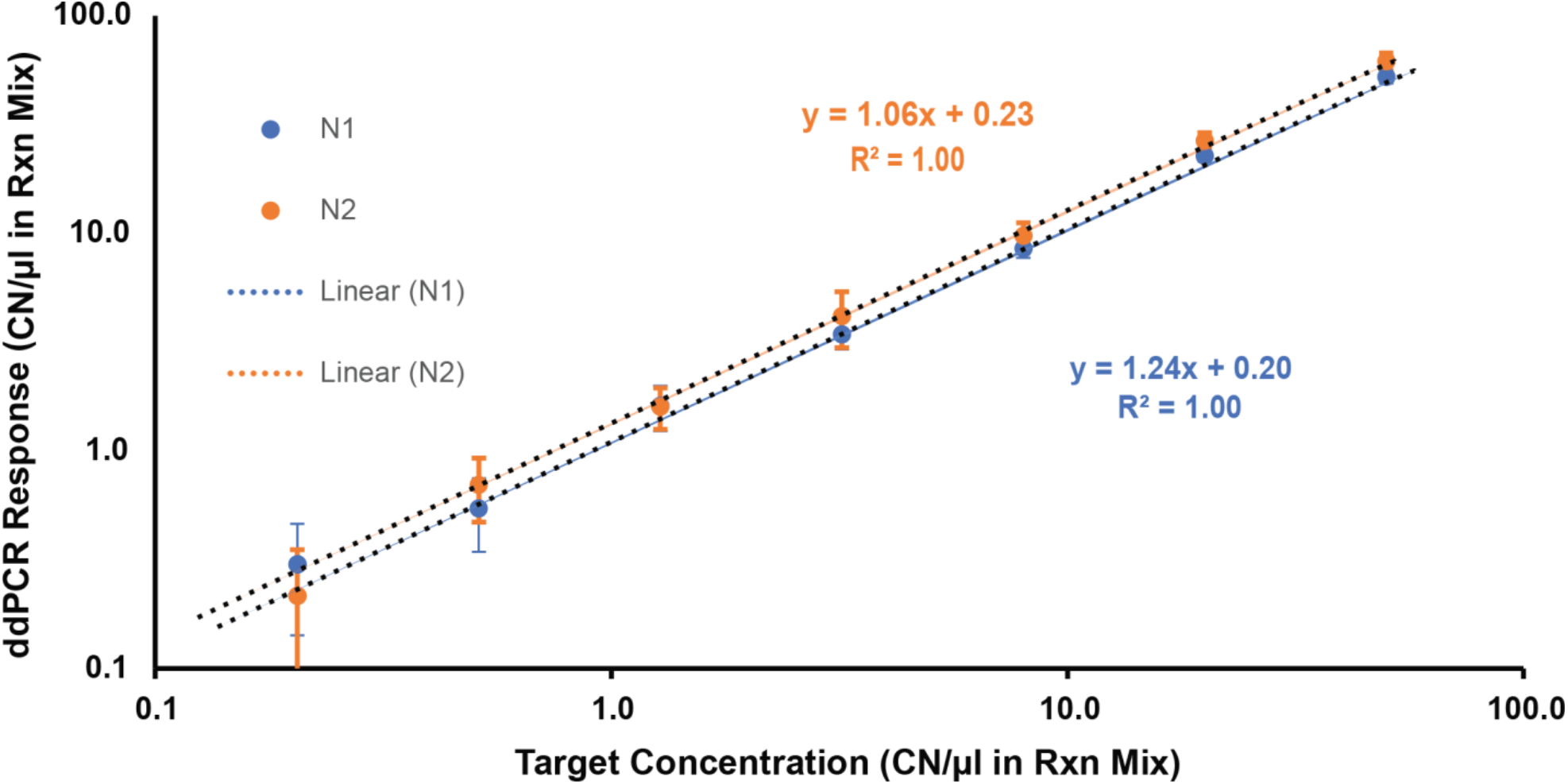
ddPCR calibration curves for N1 and N2 targets. Serial dilution of N1 and N2 standards showed calibration curves with high concordance and linearity. The limit of detection (LOD) for N1 and N2 targets are 0.06 and 0.21 CN/µl, respectively (n=2, the error bars: Standard Error).

**Supplementary Figure 3:**
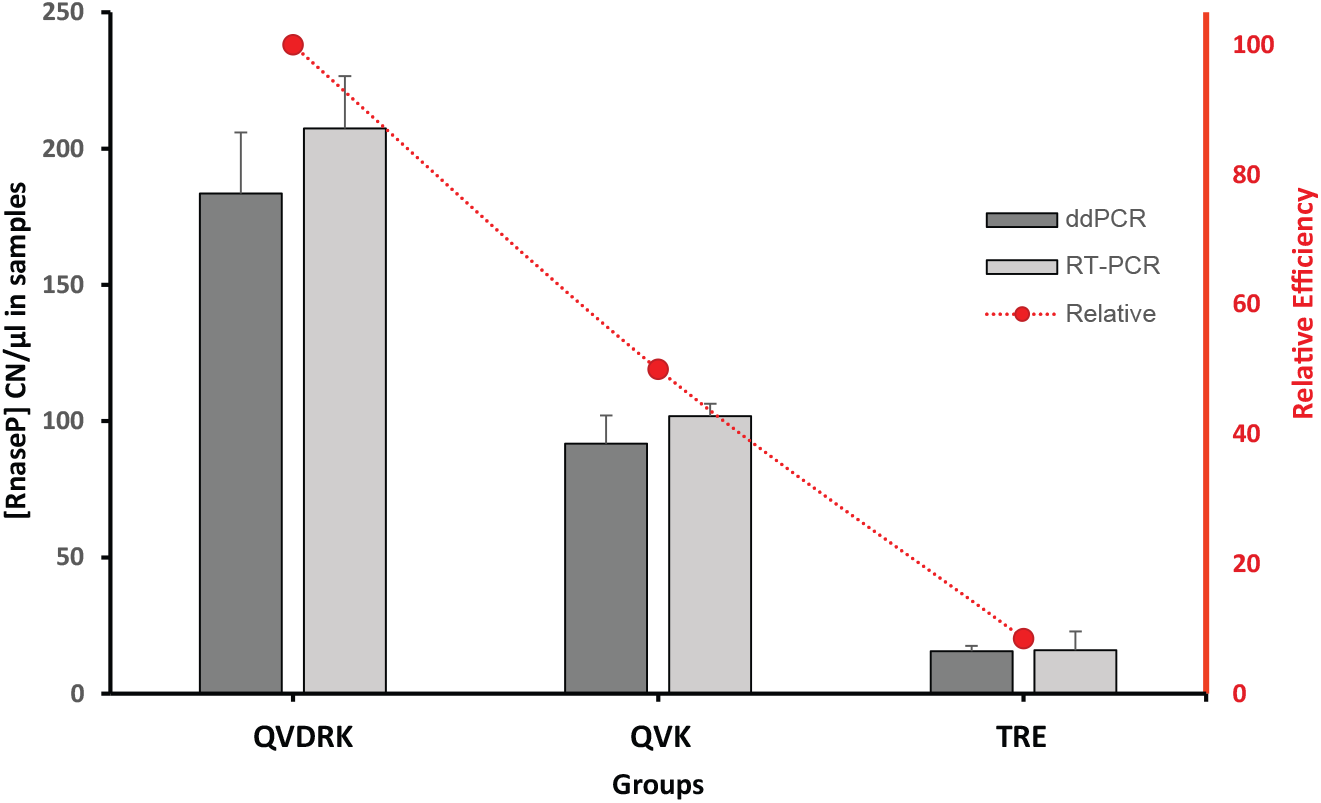
Trizol RNA Extraction Protocol (TRE) was compared with two commercial kits (i.e., QIAmp MinElute Virus DNA/RNA Spin Kit (QVDRK), QIAmp Viral RNA Mini Kit (QVK)). In this experiment, a saliva sample was aliquot into 9 vials, and each three vials were extracted by the same kit/protocol. All extracts were analyzed at the same time by RT-PCR and ddPCR to quantify RnaseP target (n=3, Error bars: Standard Deviation).

**Supplementary Figure 4:**
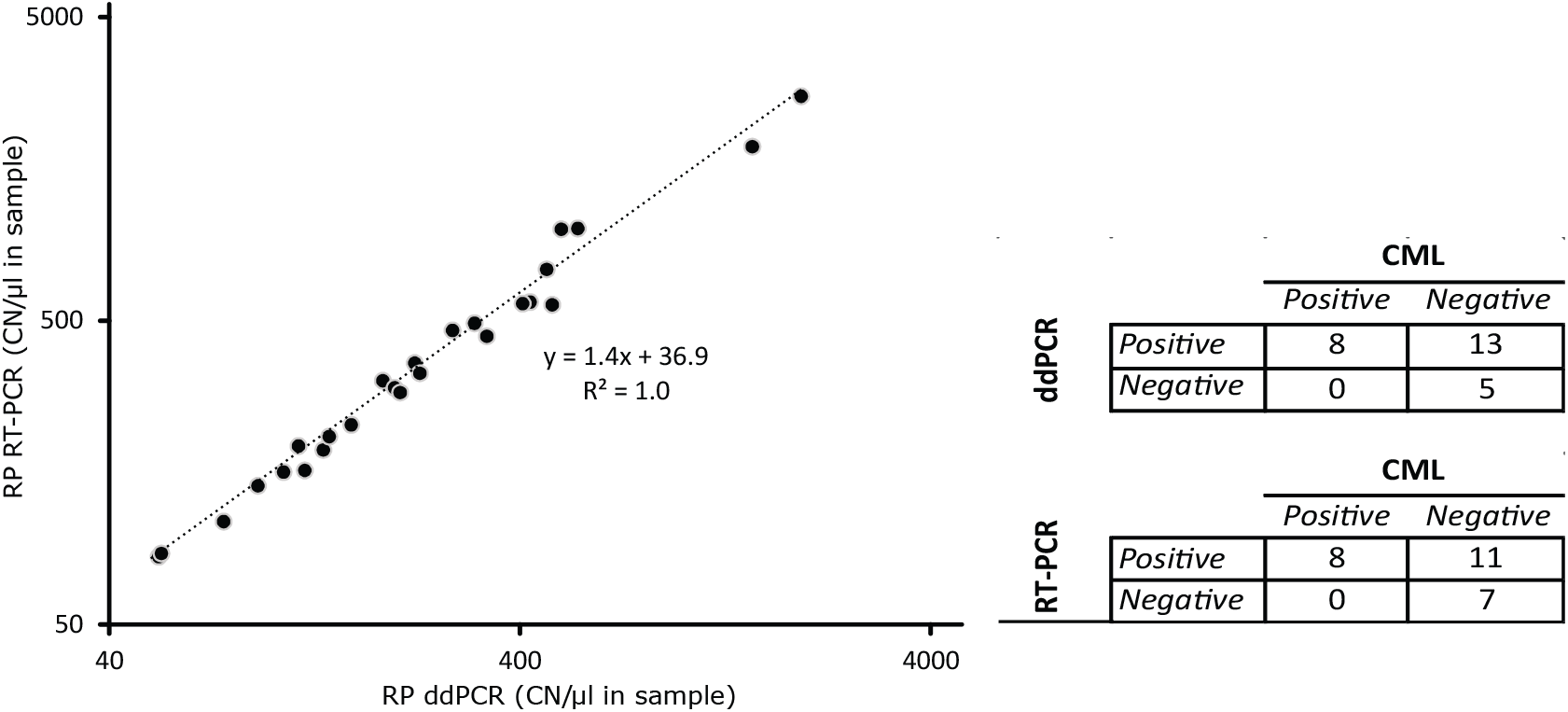
ddPCR and RT-PCR results from 26 clinically collected nasopharyngeal samples (8 positive and 18 negative by the EUA approved Xpert assay). (Left) Measurements of the RnaseP target quantified by ddPCR and RT-PCR display high correlation (Slope: 1.4 and R^2^:1.0) (n=1). (Right) The positive and negative results were compared with those of CML and showed a good concordance. Both ddPCR and RT-PCR have properly confirmed all positive results of CML. In addition, 13 and 11 CML negative samples were caught as positive by RT-PCR and ddPCR respectively.

**Supplementary Figure 5:**
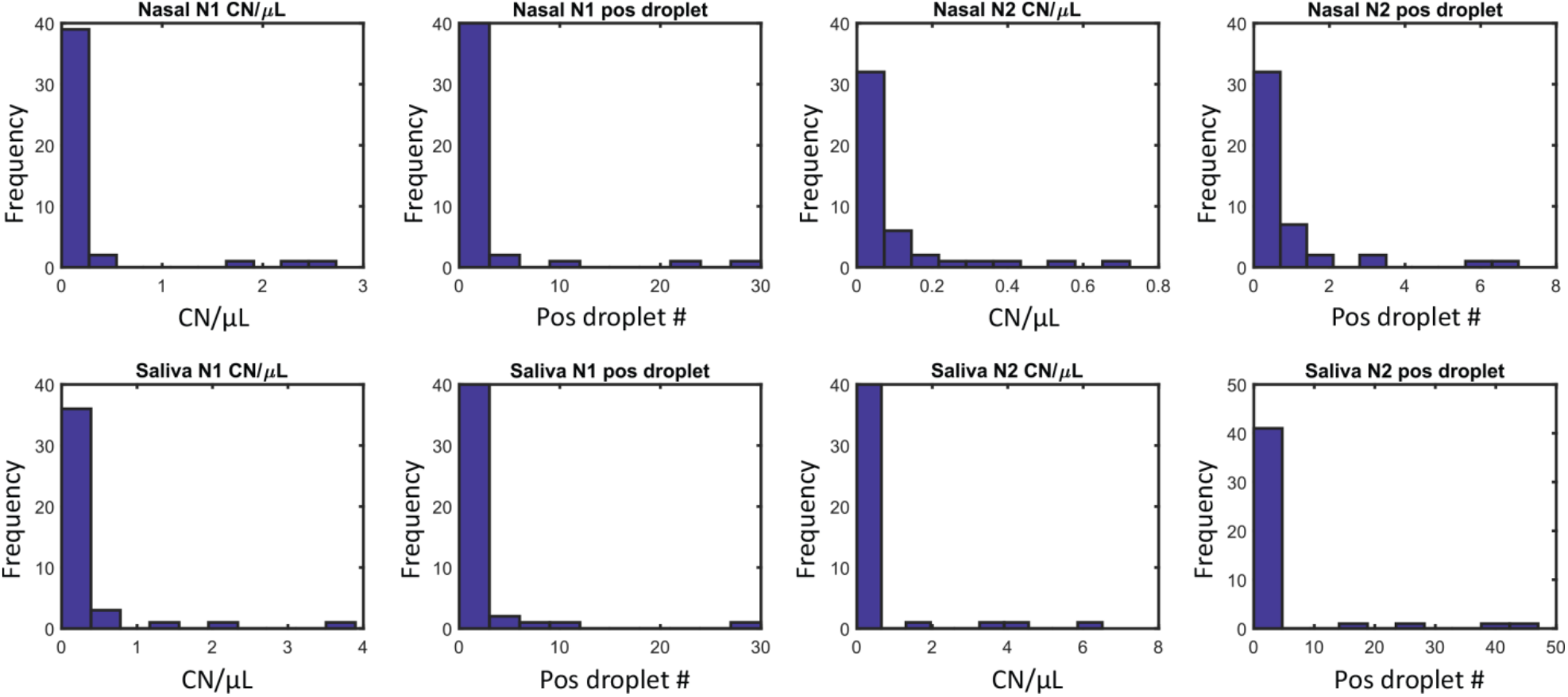
Histograms demonstrate ddPCR analysis of the specimens collected from COVID-19 negative and asymptomatic patients (n=45). Nasal swab and saliva samples were collected from symptom-free patients undergoing pre-operation screening (N = 45) and measured with digital PCR to establish an empirical threshold. The y-axis unit for all subplots are occurrences; the x-axis unit for subplots are either copy number per µL (CN/µL), or total number of positive droplets in the reaction (“pos droplet”). The thresholds were calculated as: mean+2*SD (SD: standard deviation).

**Supplementary Figure 6:**
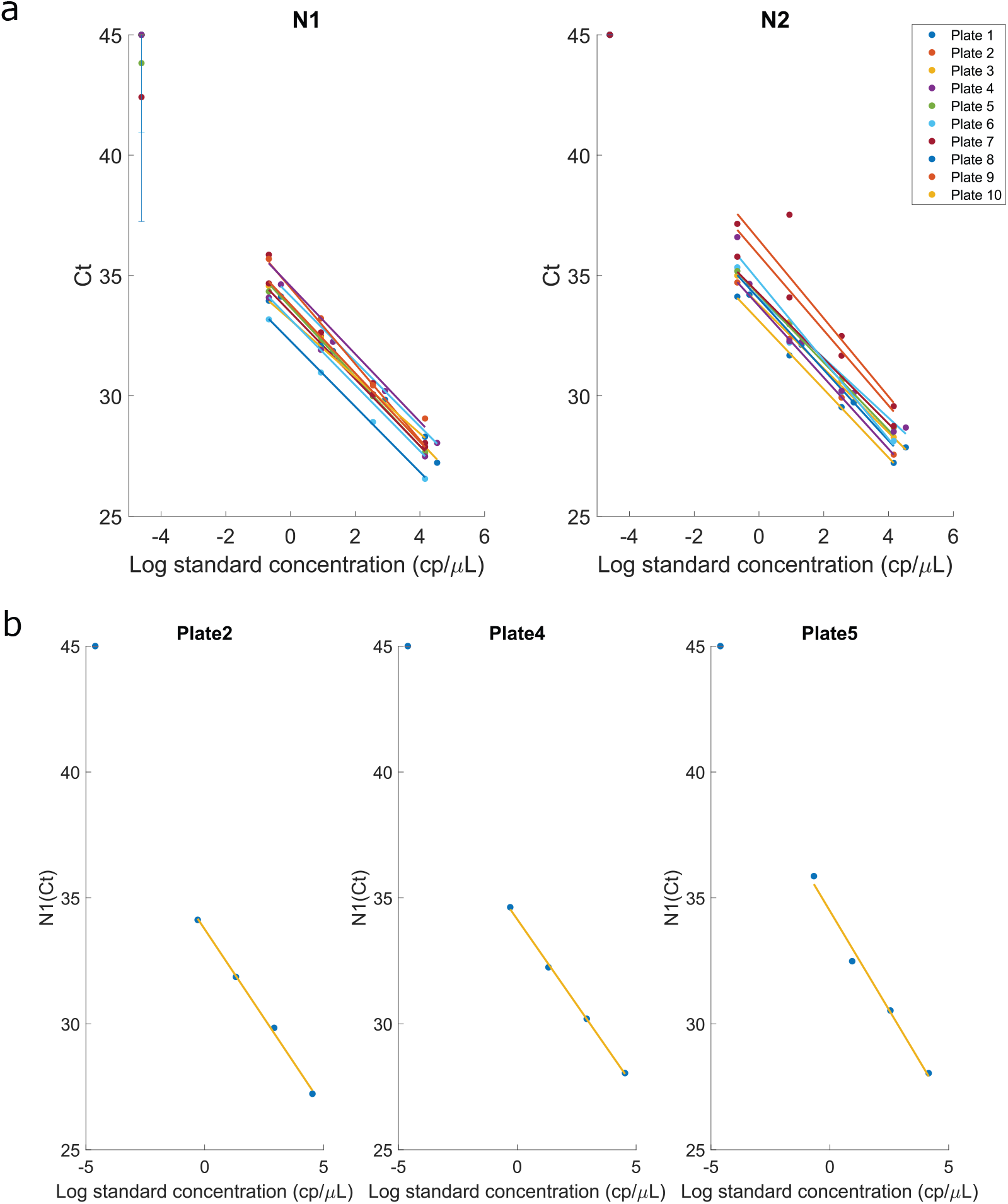
(a) RT-PCR assay validation with positive SARS-CoV-2 RNA standard dilution curve and no-template control (NTC) on all 10 plates. Standard dilution curves on three selected plates are shown in (b).

**Supplementary Figure 7:**
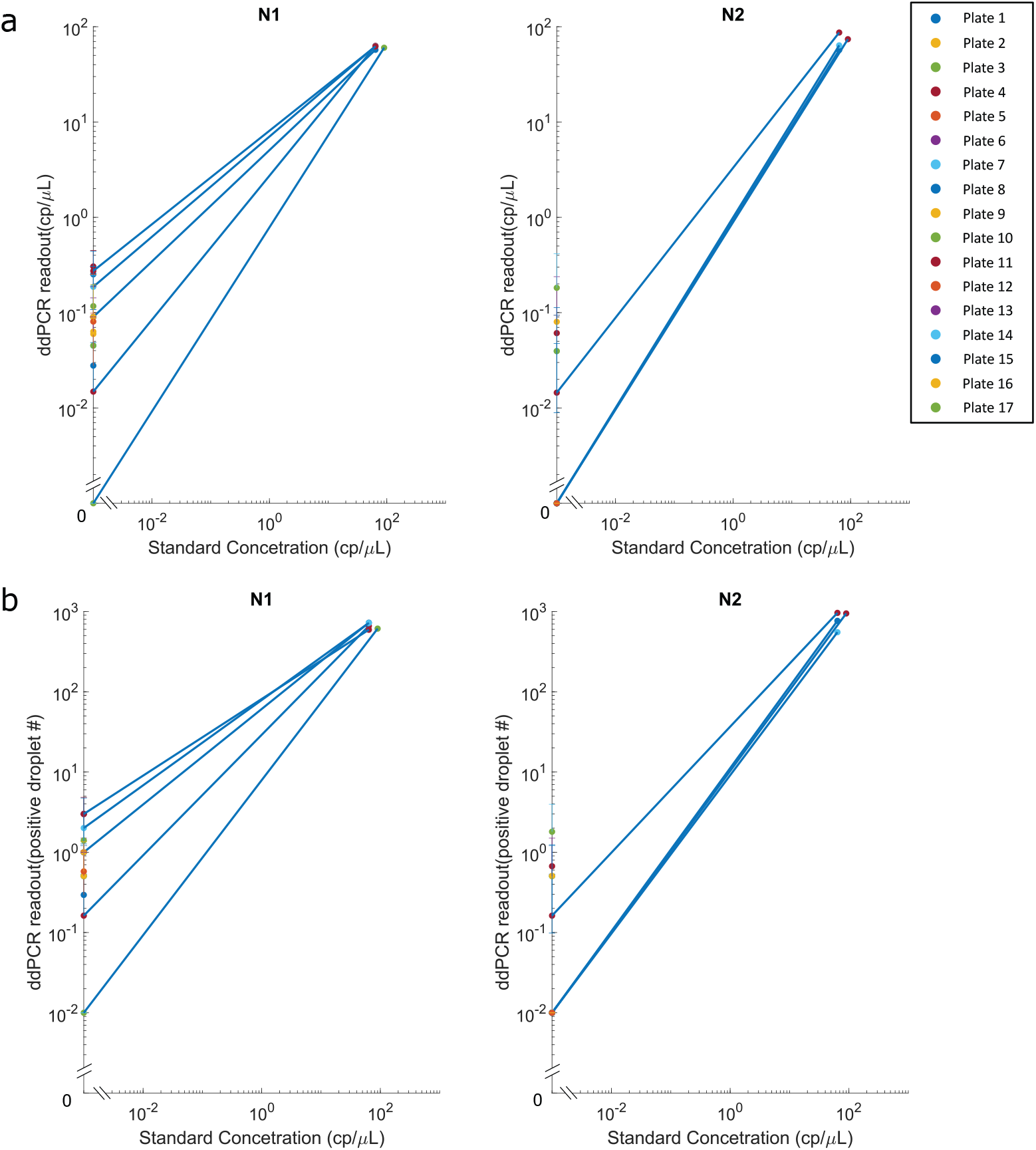
(a) ddPCR assay validation with positive SARS-CoV-2 RNA standard and no-template control (NTC) on all 17 plates, in terms of copy number per µL. (b) ddPCR assay validation with positive SARS-CoV-2 RNA standard and no-template control (NTC) on all 17 plates, in terms of positive droplet number.

**Supplementary Figure 8:**
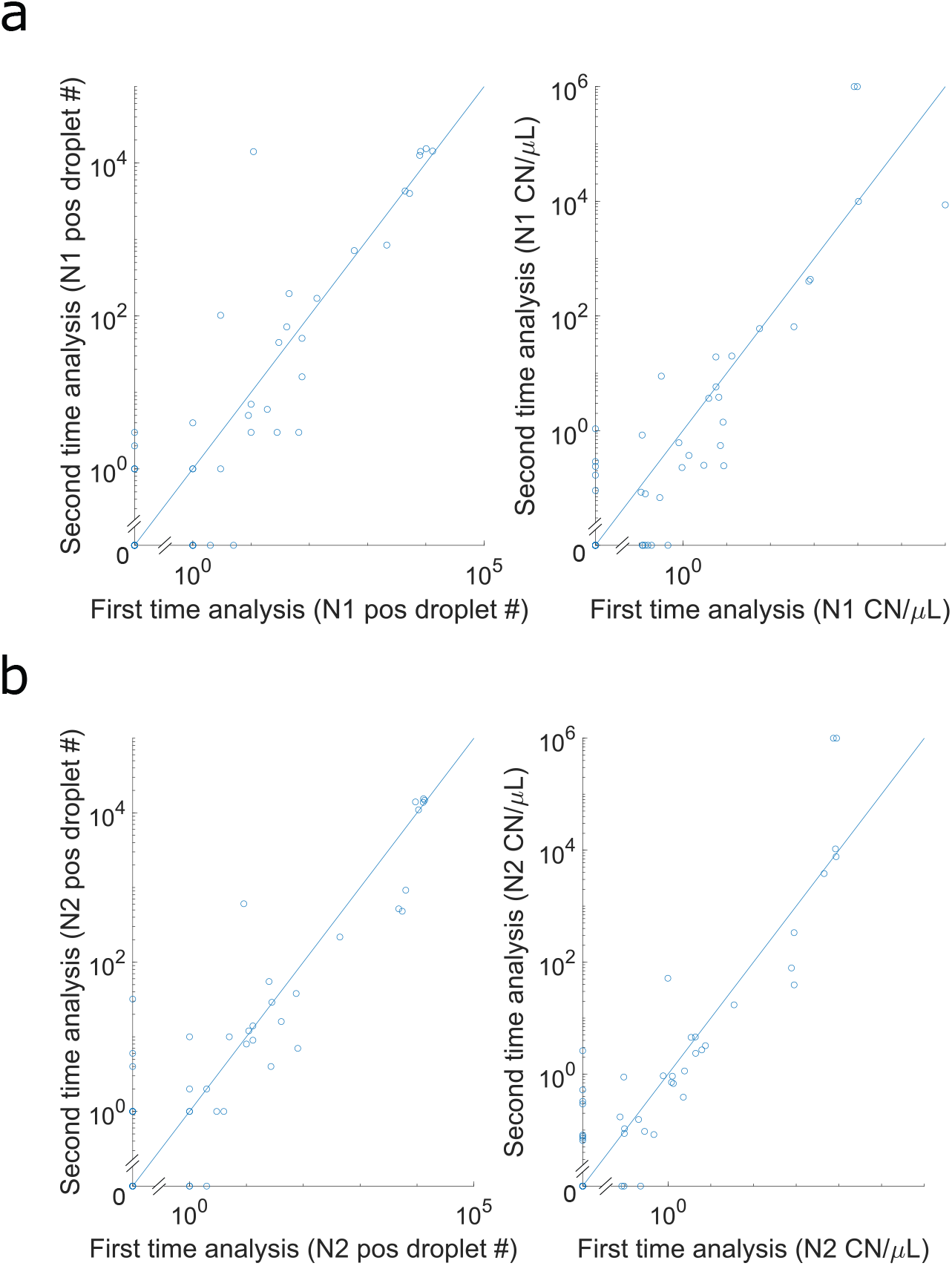
Reproducibility analysis of ddPCR assay for saliva specimens. The same extracted RNA samples (stored in -80 °C) have been re-analyzed after ∼5 months. The results indicate a high level of reproducibility. (a) The comparison of N1 target measurements of first and second analysis in terms of positive droplet number (left) and CN/µL (right); (b) The comparison of target N2 target measurements of first and second analysis in terms of positive droplet number (left) and CN/µL (right).

**Supplementary Table 1.** Comparative analysis of the EUA Xpert test performed at the microbiology laboratory, EUA TaqPath COVID-19 Combo Kit, and research laboratory-based RT-PCR/ddPCR approaches for detection of SARS-CoV-2 in pooled and diluted NP samples. Each sample was tested in duplicates.

|  | CML EUA Cepheid Xpert Xpress (Ct) |  |  |  | EUA TaqPath (Ct) |  |  |  |  |  | RL RT-PCR (Ct) |  |  |  | RL ddPCR (CN/μl/rxn mix) |  |  |  |
| --- | --- | --- | --- | --- | --- | --- | --- | --- | --- | --- | --- | --- | --- | --- | --- | --- | --- | --- |
|  | E |  | N2 |  | ORF1ab |  | N |  | S |  | N1 |  | N2 |  | N1 |  | N2 |  |
|  | Mean | SD ± | Mean | SD ± | Mean | SD ± | Mean | SD ± | Mean | SD ± | Mean | SD ± | Mean | SD ± | Mean | SD ± | Mean | SD ± |
| **1/100** | 29.8 | 0.0 | 32.5 | 0.3 | 25.0 | 0.2 | 26.1 | 0.1 | 25.2 | 0.3 | 28.7 | 0.6 | 29.5 | 1.0 | 52.6 | 18.8 | 55.0 | 32.3 |
| **1/1,000** | 33.0 | 0.3 | 36.3 | 0.4 | 28.4 | 0.0 | 29.5 | 0.1 | 28.8 | 0.2 | 31.9 | 1.0 | 33.2 | 1.1 | 5.9 | 2.9 | 6.3 | 3.9 |
| **1/10,000** | 37.8 | 1.9 | 40.2 | 1.6 | 30.6 | 1.0 | 31.7 | 0.3 | 31.8 | 0.3 | 35.4 | 0.9 | 36.6 | 0.3 | 0.6 | 0.1 | 0.6 | 0.1 |
| **1/100,000** | 0.0 | 0.0 | 42.6 | 1.4 | 33.3 | 0.7 | 34.4 | 1.2 | 36.2 | 2.8 | 38.6 | 0.1 | 39.3 | 0.9 | 0.0 | 0.0 | 0.3 | 0.0 |
| **1/1,000,000** | 0.0 | 0.0 | 0.0 | 0.0 | 0.0 | 0.0 | 0.0 | 0.0 | 0.0 | 0.0 | 42.7 | 3.9 | 45.0 | 0.0 | 0.0 | 0.0 | 0.1 | 0.1 |
| **1/10,000,000** | 0.0 | 0.0 | 0.0 | 0.0 | 0.0 | 0.0 | 0.0 | 0.0 | 0.0 | 0.0 | 45.0 | 0.0 | 45.0 | 0.0 | 0.0 | 0.0 | 0.0 | 0.0 |

RL: Research Lab: CML; Clinical Microbiology Lab

**Supplementary Table 2:** ddPCR thresholds for analysis of NS and saliva samples. Thresholds (in term of both CN/µL and total positive droplet numbers) were determined by measuring NS and saliva samples collected from COVID-19 negative patients tested at the curbside as part of their pre-operative screening (also see **Supplementary Figure 5** and **6**).

| ddPCR thresholds | Nasal |  | Saliva |  |
| --- | --- | --- | --- | --- |
|  | N1 | N2 | N1 | N2 |
| CN/μL | 1.38 | 0.38 | 1.65 | 2.95 |
| Positive droplet# | 13.27 | 3.65 | 11.51 | 23.09 |

**Supplementary Table 3:** Viral load in samples (CN/1 ml).

| Groups | Range | Nasal (Curbside) |  | Saliva (Curbside) |  | Saliva (Inpatients) |  |
| --- | --- | --- | --- | --- | --- | --- | --- |
|  | CN/ml Sample | # of Samples | Percent (%) | # of Samples | Percent (%) | # of Samples | Percent (%) |
| **SC** | 100,000 - over | 12 | 71% | 9 | 29% | 5 | 29% |
| **HL** | 10,000-100,000 | 0 | 0% | 3 | 10% | 3 | 18% |
| **ML** | 1,000-10,000 | 0 | 0% | 8 | 26% | 5 | 29% |
| **LL** | 0-1000 | 5 | 29% | 11 | 35% | 4 | 24% |

Based on viral load, samples were subclassified as follows: SC - Supercarriers, HL - High Viral Level Carriers, ML - Moderate Viral Level Carriers, LL - Low Viral Level Carriers.

**Supplementary Table 4:** List of 46 saliva samples selected for re-analysis by ddPCR.

| Patient # | CML result | RT-qPCR result | ddPCR result<br>(1st run) | ddPCR result<br>(2nd run) |
| --- | --- | --- | --- | --- |
| 1 | Negative | Negative | Negative | Negative |
| 2 | Negative | Positive | Negative | Negative |
| 3 | Negative | Negative | Negative | Negative |
| 4 | Positive | Positive | Positive | Positive |
| 5 | Negative | Negative | Negative | Negative |
| 6 | Positive | Positive | Positive | Positive |
| 7 | Negative | Negative | Negative | Negative |
| 8 | Negative | Negative | Negative | Negative |
| 9 | Negative | Positive | Negative | Negative |
| 10 | Negative | Negative | Negative | Negative |
| 11 | Negative | Negative | Positive | Positive |
| 12 | Negative | Negative | Negative | Negative |
| 13 | Negative | Negative | Positive | Negative |
| 14 | Negative | Negative | Negative | Negative |
| 15 | Negative | Negative | Negative | Negative |
| 16 | Negative | Negative | Negative | Negative |
| 17 | Negative | Negative | Positive | Positive |
| 18 | Negative | Negative | Positive | Negative |
| 19 | Negative | Negative | Positive | Negative |
| 20 | Negative | Negative | Negative | Negative |
| 21 | Positive | Positive | Positive | Positive |
| 22 | Negative | Negative | Negative | Negative |
| 23 | Negative | Positive | Positive | Positive |
| 24 | Positive | Positive | Positive | Positive |
| 25 | Positive | Positive | Positive | Positive |
| 26 | Negative | Negative | Negative | Negative |
| 27 | Negative | Negative | Negative | Negative |
| 28 | Negative | Negative | Negative | Negative |
| 29 | Negative | Negative | Negative | Negative |
| 30 | Negative | Negative | Negative | Negative |
| 31 | Negative | Negative | Negative | Negative |
| 32 | Negative | Negative | Negative | Negative |
| 33 | Negative | Negative | Negative | Negative |
| 34 | Negative | Negative | Positive | Negative |
| 35 | Negative | Negative | Negative | Negative |
| 36 | Negative | Negative | Negative | Negative |
| 37 | Positive | Positive | Positive | Positive |
| 38 | Positive | Positive | Negative | Negative |
| 39 | Positive | Positive | Positive | Positive |
| 40 | Positive | Positive | Positive | Positive |
| 41 | Negative | Positive | Positive | Positive |
| 42 | Positive | Positive | Positive | Positive |
| 43 | Negative | Positive | Positive | Positive |
| 44 | Negative | Negative | Negative | Negative |
| 45 | Positive | N.A. | Positive | Positive |
| 46 | Negative | Negative | Negative | Negative |

## Notes

### Competing Interest Statement

The authors have declared no competing interest.

### Funding Statement

This study was supported in part by U.S. National Institutes of Health (NIH) grants R01GM127527 to S.T., R01DE027809 E. I., and R01DE028674 to N. A., and Paul. G. Allen Distinguished Investigator Award to S.T. We thank The University of Chicago Pritzker School of Molecular Engineering, and the Biological Sciences Division, for financially supporting this study.

